# Impact of a public policy restricting staff mobility between long-term care homes in Ontario, Canada during the COVID-19 pandemic

**DOI:** 10.1101/2020.11.17.20231498

**Authors:** Aaron Jones, Alexander G. Watts, Salah Uddin Khan, Jack Forsyth, Kevin A. Brown, Andrew P. Costa, Isaac I. Bogoch, Nathan M. Stall

## Abstract

**Objectives:** To assess changes in the mobility of staff between long-term care homes in Ontario, Canada before and after enactment of public policy restricting staff from working at multiple homes.

**Design:** Pre-post observational study.

**Setting and Participants:** 623 long-term cares homes in Ontario, Canada between March 2020 and June 2020.

**Methods:** We used anonymized mobile device location data to approximate connectivity between all 623 long-term care homes in Ontario during the 7 weeks before (March 1 – April 21) and after (April 22 – June 13) the policy restricting staff movement was implemented. We visualized connectivity between long-term care homes in Ontario using an undirected network and calculated the number of homes that had a connection with another long-term care home and the average number of connections per home in each period. We calculated the relative difference in these mobility metrics between the two time periods and compared within-home changes using McNemar’s test and the Wilcoxon rank-sum test.

**Results:** In the period preceding restrictions, 266 (42.7%) long-term care homes had a connection with at least one other home, compared to 79 (12.7%) homes during the period after restrictions, a drop of 70.3% (p <0.001). The average number of connections in the before period was 3.90 compared to 0.77 in after period, a drop of 80.3% (p < 0.001). In both periods, mobility between long-term care homes was higher in homes located in larger communities, those with higher bed counts, and those part of a large chain.

**Conclusions and Implications:** Mobility between long-term care homes in Ontario fell sharply after an emergency order by the Ontario government limiting long-term care staff to a single home, though some mobility persisted. Reducing this residual mobility should be a focus of efforts to reduce risk within the long-term care sector during the COVID-19 pandemic.

## Introduction

Worldwide, residents of long-term care (LTC) homes have been disproportionately affected by the COVID-19 pandemic.^1^ In Canada, nearly 80% of all COVID-19 deaths have occurred among LTC home residents, the highest known proportion among developed countries.^2,3^ Residents of LTC homes are at disproportionately increased risk of both COVID-19 infection and poor COVID-19 outcomes, as a result of congregate living, advanced age, frailty and multimorbidity.^4^

Early in the pandemic, healthcare workers were identified as important and unknowing vectors for importation of COVID-19 into LTC homes, with movement of staff between LTC homes and other healthcare settings being key contributors to the spread of COVID-19.^5–7^ Staff in LTC homes are frequently employed on a part-time or casual basis, requiring individuals to work in multiple homes or health care settings to earn a living wage.^8,9^

When COVID-19 outbreaks intensified in LTC homes during the first months of the pandemic, numerous jurisdictions enacted policies to limit the movement of LTC staff between multiple homes.^10–12^ On April 22^nd^, 2020, Canada’s most populous province of Ontario implemented an order that prevented staff from working in more than one home. Recognizing that existing staffing shortages were exacerbated by the COVID-19 pandemic,^13^ the order did not apply to temporary agency workers or other contract staff, in order to ensure LTC homes had an availability of staff to work on an emergency basis.^14,15^ We used anonymized mobile GPS location data to examine the impact of this order on mobility between LTC homes.

## Methods

### Setting and Study Design

We conducted a pre-post observational study of mobility between LTC homes in Canada’s most populous province of Ontario (population approximately 15 million residents). Medical and personal care in LTC homes are covered by Ontario’s universal and publicly funded health insurance plan, with residents responsible for an accommodation copayment. Currently, there are over 70,000 residents in 623 long-term care homes in Ontario.^16^

### Data sources

We obtained all data for this study as part of the COVID-19 Ontario Census Modelling Table. The Modelling Table is sponsored by the Ontario Ministry of Health, Ontario Health, and Public Health Ontario and is an ad-hoc and voluntary group of senior decision-makers and scientists tasked with creating credible consensus estimates of the impact of COVID-19. We used anonymized, population-aggregated, near-real-time, mobile device GPS location data provided by Veraset (Veraset, San Francisco, CA), a data-as-a-service vendor. Verasat aggregates location data across several apps on both Apple and Android platforms after the user has consented to use of their anonymized data, and has been previously used in research.^7,17^ Data on long-term care home characteristics and COVID-19 outbreaks were obtained from the Ontario Ministries of Health and Long-Term Care. The study was approved by the Research Ethics Board of (masked).

### Emergency order

On April 15, 2020 the Government of Ontario announced an emergency order restricting employees of LTC homes from working in more than one LTC home, congregate care or healthcare setting within a 14- day period. The order came into effect on April 22, 2020, but did not apply to temporary agency staff or other contract staff (who are not employees of LTC homes), in order to ensure LTC homes had an availability of staff to work on an emergency basis location^18^.

### Mobility between long-term care homes

We evaluated mobility between LTC homes in two time periods before and after implementation of the emergency order: March 1 - April 21, 2020 (the “before” period) and April 22 - June 13, 2020 (the “after” period). We used Statistics Canada’s Open Database of Buildings to define home location and footprint^19,20^. For homes where there was no entry in the database, we defaulted to a 50-meter buffer around a home’s latitude-longitude coordinates. A visit to a home occurred if a device had GPS check-ins within a LTC home’s boundary in two consecutive half-hour time blocks with a minimum total elapsed time of thirty minutes, thereby selecting visits that are likely to be staff entering homes rather than transient visits due to deliveries, etc. A connection between two homes was recorded if the same device had visits to two separate homes within 14 days. Of note, the Province of Ontario restricted visitors to LTC homes from March 14-June 18, 2020 with the exception of end-of-life compassionate visits, meaning that mobility devices captured within our study period would very likely be limited to residents and staff.

We calculated mobility metrics for each LTC home based on the connections in which it was the destination home, i.e. a device entered the home after being detected in a different LTC home in the previous 14 days. For each home we calculated whether there was any connection with another LTC home and the total count of connections with other homes.

### Analysis

We constructed an undirected network connectivity diagrams for periods before and after restrictions were implemented. We identified homes in COVID-19 outbreak (≥ 1 resident or staff case) during any part of each period on the diagrams. For each of our network metrics, we calculated the average for the before and after periods, and the relative difference (expressed as a percentage) between the two time periods. We reported these measures overall and by home size (number of licensed beds), community size (population), for-profit status (for-profit, non-profit or municipal), and chain size (single home, small chain [2-19], or large chain [≥20]). Homes owned by municipalities were not considered part of a chain. We performed within-home comparisons of the mobility metrics between the periods using McNemar’s test and the Wilcoxon signed-rank test. Analysis was done using R 4.0.0^21,22^.

## Results

We identified 5,640 unique mobile devices that entered at least one of Ontario’s 623 LTC homes in the 7 weeks before enactment of an emergency order restricting staff mobility between LTC homes (March 1 - April 21, 2020) and 3,792 unique devices that entered in the 7 weeks after enactment of the order (April 22 - June 13, 2020). Prior to the emergency order, 266 (42.7%) LTC homes had a connection with at least one other LTC home, compared to 79 (12.7%) homes following the order, a drop of 70.3% (p <0.001) (Figure 1 and Table 1). The average number of connections was 3.90 in the time period before the order and 0.77 in the time period following the order, corresponding to a drop of 80.3% (p < 0.001). Mobility between LTC homes was higher in homes located in larger communities, in those with higher bed counts, and those part of a large chain. During the after period, 72 (30.2%) of homes with ≥ 200 beds still had at least one connection compared to 65 (1.5%) of the homes with ≤ 20 beds and 145 (17.9%) of the large chain homes had a connection compared to 268 (9.3%) of the homes that were not part of a chain. There were 149 (23.9%) homes in outbreak at any point during the before period, and 292 (46.9%) homes in outbreak at any point during the after period.

**Table 1:** Mobility between Ontario long-term care homes before (March 1-April 21, 2020) and after (April 22-June 13, 2020) implementation of an emergency order limiting staff to working at one long-term care home or healthcare setting

|  | n | % Homes with a Connection |  |  |  | Average # of Connections per home |  |  |  |
| --- | --- | --- | --- | --- | --- | --- | --- | --- | --- |
|  |  | Before | After | % diff | p<sup>1</sup> | Before | After | % diff | p<sup>2</sup> |
| Overall | 623 | 42.7% | 12.7% | -70.3% | <0.001 | 3.90 | 0.77 | -80.3% | <0.001 |
| Profit Status |  |  |  |  |  |  |  |  |  |
| For-profit | 360 | 43.1% | 14.7% | -65.9% | <0.001 | 3.94 | 0.94 | -76.1% | <0.001 |
| Not-for-profit | 162 | 46.3% | 11.1% | -76.0% | <0.001 | 4.02 | 0.50 | -87.6% | <0.001 |
| Municipal | 101 | 35.6% | 7.9% | -77.8% | <0.001 | 3.47 | 0.60 | -82.7% | <0.001 |
| Chain Type |  |  |  |  |  |  |  |  |  |
| Large (≥20) | 145 | 50.3% | 17.9% | -64.4% | <0.001 | 5.16 | 1.50 | -70.9% | <0.001 |
| Small (2-19) | 210 | 44.3% | 13.3% | -70.0% | <0.001 | 3.95 | 0.58 | -85.3% | <0.001 |
| Single home | 268 | 37.3% | 9.3% | -75.1% | <0.001 | 3.15 | 0.53 | -83.2% | <0.001 |
| Home Size |  |  |  |  |  |  |  |  |  |
| ≥200 beds | 76 | 67.1% | 30.2% | -55.0% | <0.001 | 8.08 | 2.58 | -68.1% | <0.001 |
| 100-199 beds | 201 | 52.0% | 15.3% | -70.6% | <0.001 | 5.23 | 0.91 | -82.6% | <0.001 |
| 51-100 beds | 281 | 27.8% | 6.0% | -78.4% | <0.001 | 1.53 | 0.13 | -91.5% | <0.001 |
| ≤ 50 beds | 65 | 20.0% | 1.5% | -92.5% | <0.001 | 0.47 | 0.03 | -93.6% | <0.001 |
| Community Size (population) |  |  |  |  |  |  |  |  |  |
| ≥500,000 | 255 | 65.9% | 21.6% | -67.2% | <0.001 | 7.56 | 1.65 | -78.2% | <0.001 |
| 100,00-499,999 | 225 | 32.4% | 8.0% | -75.3% | <0.001 | 1.83 | 0.24 | -86.9% | <0.001 |
| <10,000 (rural) | 143 | 17.5% | 3.5% | -80.0% | <0.001 | 0.53 | 0.06 | -88.7% | <0.001 |

1. McNemar's test
2. Wilcoxon rank-sum test

**Figure 1:**
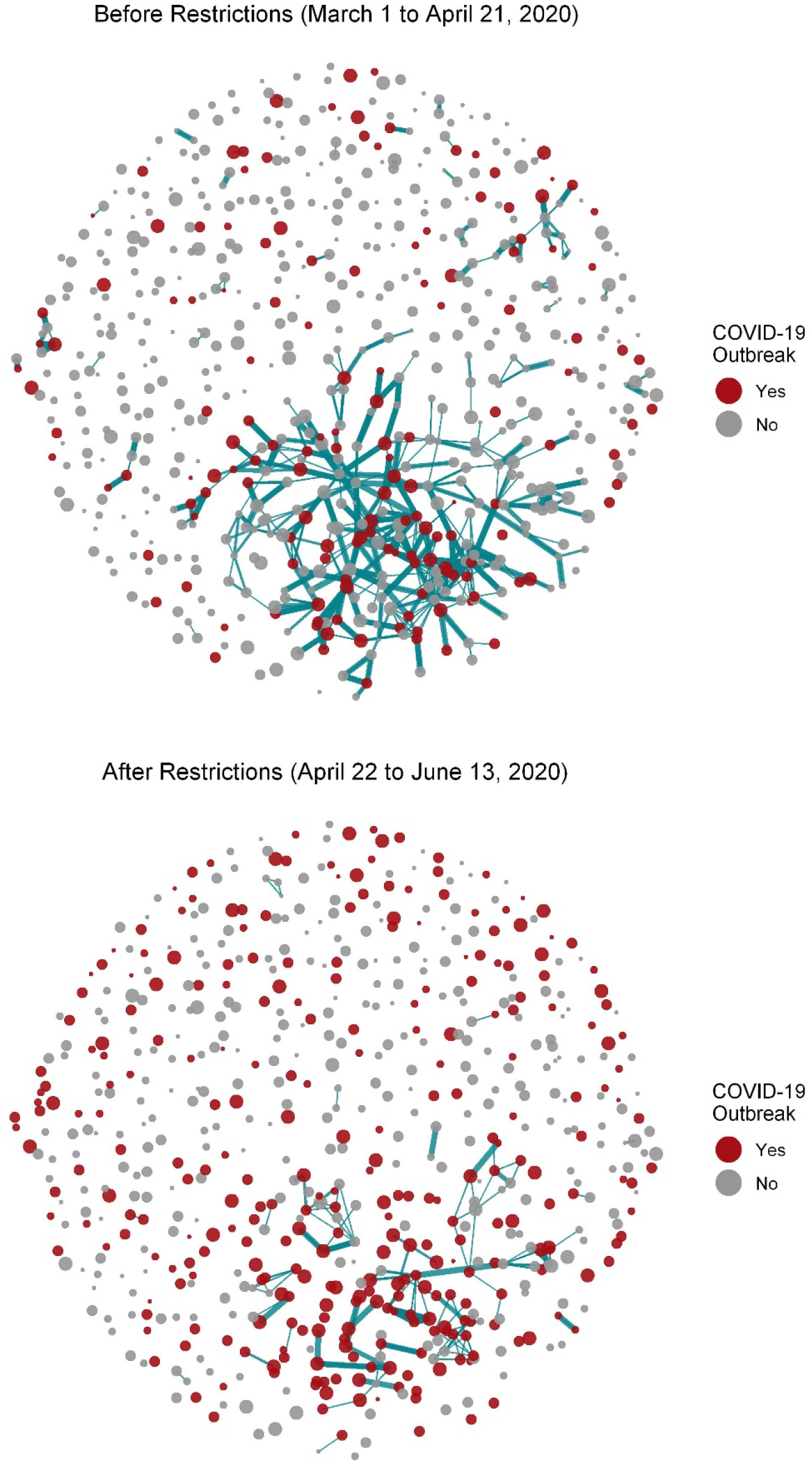
Network connectivity diagrams before and after emergency order limiting staff to working at one long-term care home or healthcare setting.

## Discussion

Mobility between LTC homes in the Province of Ontario dropped sharply after implementation of an emergency order restricting staff to work a single home. The number of LTC homes with any connection to another home fell by 70.3% while the total number of connections dropped by 80.3%. In spite of this, 12.7% of all Ontario LTC homes still had connections following enactment of the emergency order.

Staff mobility between LTC homes has been identified as an important vector for importation of COVID- 19 into and spread between homes, and many jurisdictions have therefore restricted or limited workers from working in multiple healthcare settings. Ontario’s emergency order came into force three and half weeks after the first reported COVID-19 outbreak in a LTC home (March 29 to April 22, 2020), at which point nearly 150 homes had experienced an outbreak.^7^ Other Canadian provinces like British Columbia responded somewhat faster, limiting mobility in just under three weeks after their first reported outbreak (March 7 to March 26, 2020)^13^, at which point only 9 homes were in outbreak.^24^ Given the robust drop in mobility we observed in our study, and other research linking higher staff mobility to increased COVID-19 incidence in LTC homes, delays in limiting mobility may have resulted in additional cases and deaths among LTC residents and staff.^7,25^

The ongoing connectivity of 79 (12.7%) of Ontario’s 623 LTC homes following implementation of the order merits serious consideration, especially as decision-makers and the LTC sector grapple with the intensifying second wave of the COVID-19 pandemic. This connectivity could represent non-compliance with the provincial order or movement of individuals not covered by the order such as physicians who work at multiple homes. The persistent connectivity may represent Ontario exempting temporary agency workers and contract staff from the ban on working at more than one home or healthcare setting. Any residual mobility remains a potential vector for the importation of COVID-19 into LTC homes and closing the loophole on the emergency order could reduce risk in the sector during successive waves of the pandemic. Doing so would require addressing chronic staffing shortages, by accelerating the training of LTC workers and by promoting retention in the field by ensuring all workers have fair and full-time pay with benefits and a career ladder to support their advancement.^26^

### Limitations

The mobility location data acquired from Veraset relies on GPS check-ins from several mobile phone applications. As such, individuals who do not use these applications or do not consent to sharing their data will not be captured. While this will underestimate absolute measures of mobility, the relative differences calculated between the periods are robust. Furthermore, a dearth in geospatial footprint data per facility limited the spatial accuracy in detectability of devices by a generalized 50 metre buffer although this sampling error was consistent across all facilities. Finally, we were not able to assess the temporal association between staff mobility and COVID-19 outbreaks in LTC homes as this would require longitudinal mobility data on a granular level.

## Conclusions and Implications

The emergency order by the Ontario government limiting LTC staff to a single home sharply reduced mobility between LTC homes. The residual mobility between 79 (12.7%) of Ontario’s 623 LTC homes remains an potential vector for the importation of COVID-19 into LTC homes and should be the focus of efforts to reduce risk within the LTC sector during the COVID-19 pandemic.

## Data Availability

The data from this study is not publicly available. The underlying analytic code are available from the authors upon request.

